# A COVID-19 Community Vulnerability Index to drive precision policy in the US

**DOI:** 10.1101/2021.05.19.21257455

**Authors:** Peter Smittenaar, Nicholas Stewart, Staci Sutermaster, Lindsay Coome, Aaron Dibner-Dunlap, Mokshada Jain, Yael Caplan, Christine Campigotto, Sema K. Sgaier

## Abstract

**Background:** In April 2020 we released the US COVID-19 Community Vulnerability Index (CCVI) to bring to life vulnerability to health, economic, and social impact of COVID-19 at the state, county, and census tract level. Here we describe the methodology, how vulnerability is distributed across the U.S., and assess the impact on vulnerable communities over the first year of the pandemic.

**Methods:** The index combines 40 indicators into seven themes, drawing on both public and proprietary data. We associate timeseries of COVID-19 cases, deaths, test site access, and rental arrears with vulnerability.

**Results:** Although overall COVID-19 vulnerability is concentrated in the South, the seven underlying themes show substantial spatial variability. As of May 13, 2021, the top-third of vulnerable counties have seen 21% more cases and 47% more deaths than the bottom-third of vulnerable counties, despite receiving 27% fewer tests (adjusted for population). Individual vulnerability themes vary over time in their relationship with mortality as the virus swept across the country. Over 20% of households in the top vulnerability tercile have fallen behind on rent. Poorer test site access for rural vulnerable populations early in the pandemic has since been alleviated.

**Conclusion:** The CCVI captures greater risk of health and economic impact. It has enjoyed widespread use in response planning, and we share lessons learned about developing a data-driven tool in the midst of a fast-moving pandemic. The CCVI and an interactive data explorer are available at precisionforcovid.org/ccvi.

What is already known on this topic

- Various communities across the United States will experience the adverse effects of public health crises to different degrees of severity.
- Composite indicators, such as the CDC Social Vulnerability Index, have proven to be valuable to policymakers by turning complex data sets into easily digestible and actionable information. However, the indicators within the Social Vulnerability Index do not fully contextualize the negative impacts spurred by the current pandemic.

What this study adds

- The U.S. COVID-19 Community Vulnerability Index captures vulnerabilities spanning health, social, and economic dimensions that have been felt by every community in the US differently.
- Vulnerable populations have experienced more cases and deaths, higher unemployment, and a lack of access to critical support such as testing sites.
- Precision policies targeting vulnerable populations need to be designed and enacted to decrease the gap in negative consequences experienced in this and future pandemics, and the COVID-19 Community Vulnerability Index is a tool to highlight where and why these inequities occur.

## Introduction

On January 21st, the first case of COVID-19 was detected in the United States (US). By mid-March, community transmission had led to spread across the country with an estimated 100,000-200,000 people walking around infected (1). This triggered a set of drastic policies that shut down substantial parts of the economy, education system, and society more broadly. This approach was exceptionally blunt, causing widespread economic damage (2), job losses, and secondary health impacts such as delayed cancer diagnoses and a rise in poor mental health (3, 4). These impacts were by no means equally distributed across the population, and vulnerable communities bore the brunt of the negative consequences (5), with more mental health issues in vulnerable communities (3) and impact on employment and earnings (2, 6). Similar to previous pandemics (7), COVID-19 has exacerbated health and economic inequalities.

In March 2020, we started exploring how to provide policymakers with tools to protect these vulnerable communities across the US, both in terms of their health and their social and economic wellbeing. Supporting communities using limited resources requires a precision approach (8), meaning the application of policies and interventions that take into account local social, economic, and health conditions (9). It was known before COVID-19 that many socioeconomic and environmental factors mediate the impact of a pandemic (10–12), and preliminary evidence from China and the United States was pointing to specific risk factors for poor outcomes such as old age (13). There was a clear need to quickly map which communities would be most vulnerable and for what reason(s), such that policymakers could proactively prioritize limited resources to the right communities. Looking beyond the initial wave of infections, such a granular map would continue to provide key context for other policies such as re-opening of businesses, targeting health and nutrition services, providing financial support, and ensuring equitable vaccine distribution. While several pre-COVID-19 public health indices were available (14–17), all lacked factors critical to COVID-19, geographic granularity for an effective response, or both.

To fill this gap, we constructed the COVID-19 Community Vulnerability Index (CCVI) to predict which US communities would be less resilient to the COVID-19 pandemic. We defined vulnerability as a limited ability to mitigate, treat, and delay transmission of the virus and to withstand its secondary effects on health, economic and social outcomes. By definition, vulnerability is a multidimensional construct, and we designed a modular index to reflect this. The CCVI has since been recognized by the Centers for Disease Control and Prevention as a valuable tool in COVID-19 research and pandemic response planning (18, 19).

In this paper we describe the methodology and data sources behind the index. We present the results of this work, including the CCVI itself and secondary data showing the disproportionate impact of COVID-19 on the health and economic wellbeing of vulnerable communities.

## Methodology

### Building on existing indices using the latest evidence

The CCVI incorporated a large range of indicators in order to capture the many facets of vulnerability. We started with a validated index, the CDC’s Social Vulnerability Index (SVI), that was readily available at the census tract level (15, 20). The SVI contains four themes - socioeconomic status, household composition and disability, minority status and language, and housing type and transportation - that capture populations disproportionately affected by disasters (15, 20). The SVI was not designed specifically for the COVID-19 pandemic. In particular, it did not include a range of risk factors for poor clinical outcomes of the virus, living and working conditions that lead to adverse and disproportionate COVID-related outcomes, or metrics of health system capacity.

We released an initial version of the index in April 2020, which added epidemiological and health system themes to an otherwise unchanged SVI. We released an updated version of the index in November 2020, which is the index described in the remainder of this paper. The CCVI consists of seven themes, with the first three reflecting a condensed version of the SVI, and the remaining four reflecting specific COVID-19 themes (Figure 1). Each theme consists of indicators available at the census tract, county, or state level, described in detail in Appendix 1. If an indicator was unavailable at the census tract, the value for the parent region was used.

**Figure 1:**
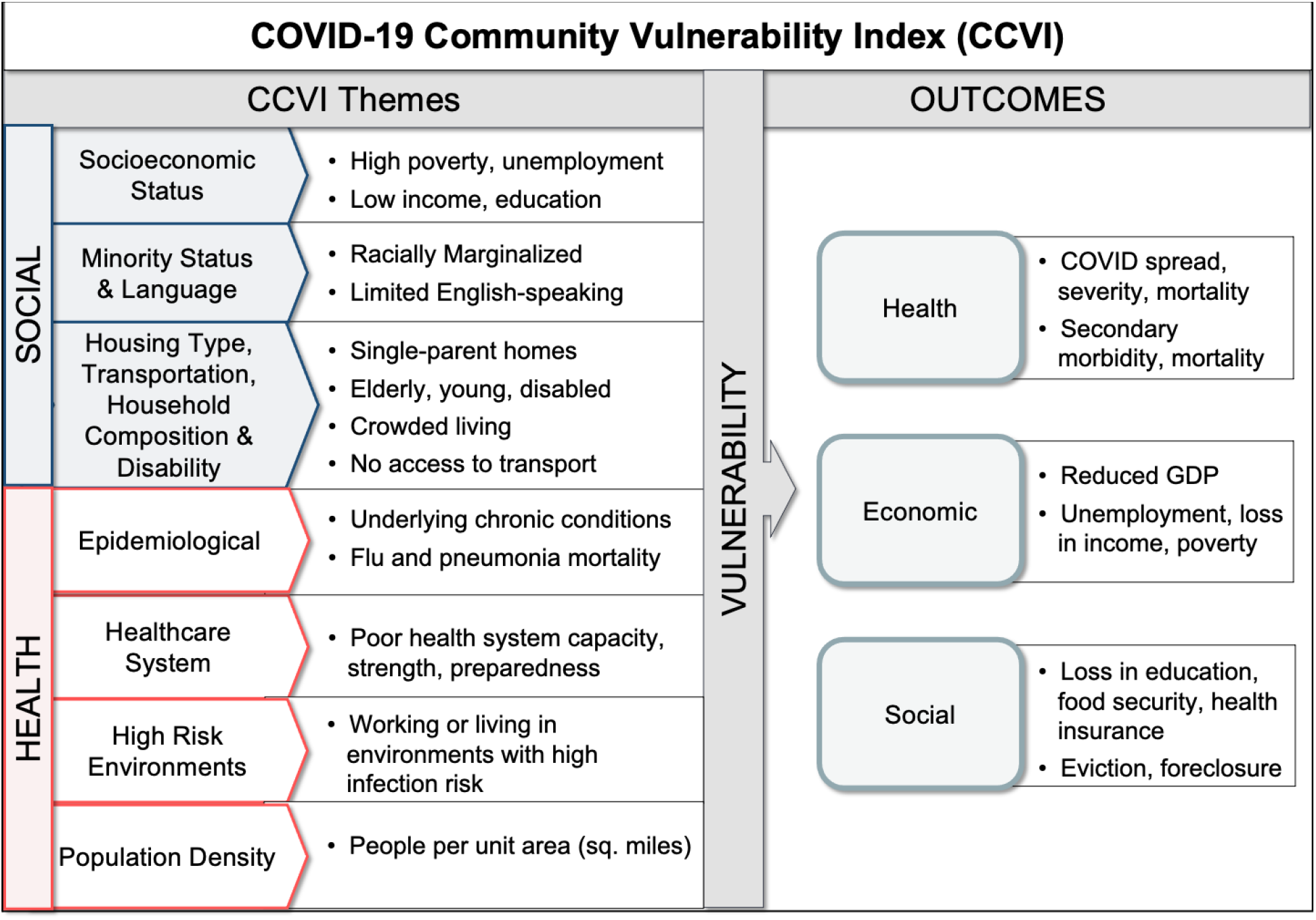
CCVI themes and subthemes, and outcome measures. The themes of the index cover a variety of underlying factors that drive vulnerability to a wide range of adverse pandemic outcomes, some of which are listed on the right. The raw indicators are enumerated in Appendix 1; a referenced rationale describing relationships between themes and outcomes is shown in Appendix 2.

To decide on the themes, we evaluated the COVID-19 literature on aggravating factors for poor health, social, and economic outcomes up to November 2020, and discussed the index with subject-matter experts. Appendix 2 captures the evidence base underpinning each of the themes, and Appendix 34 describes how the indicators were statistically aggregated into subthemes, themes, and an overall index score.

### Statistical analysis of the index with secondary data

Dynamic datasets related to COVID-19 were collected for analysis against the index and its constituent themes. Confirmed COVID-19 cases and death counts were obtained on March 2, 2021 from the Johns Hopkins University Center for Systems Science and Engineering, which collects and reports COVID-19 data for each US county (21). We calculated case and death rates as the average number of newly confirmed cases and deaths over a 7-day period. COVID-19 testing location data were compiled from GISCorps (22). Minimum travel distance to testing location was calculated as Manhattan (city-block) Haversine distances between a population-weighted census tract centroid and the nearest test site; county-level estimates were created using the population-weighted census tract mean.

We examined the association between vulnerability and rental arrears at the state level through the U.S. Census Bureau’s Household Pulse Survey (23), a repeated cross-section instrument which measures personal and household responses to the pandemic, including rental payment behavior. We judged a renting household to be in arrears if they responded negatively to the question: “Are you currently up to date on your rent payments?” We estimated state-level impact of the CCVI on rental arrears using the generalized least squares method using household replication weights in the *survey* package in R (24). Reported p-values correspond to the coefficient on the CCVI predictor.

We performed principal components analysis to understand the shared properties of the CCVI’s underlying themes using the FactoMineR package in R (25), with the corresponding loadings reported in a distance biplot. An analysis of the similarities and differences between the CCVI and SVI is presented along with a regression of each index against several negative outcomes related to the COVID-19 pandemic. To do this, we conducted bivariate regressions with the index as the predictor and state as a fixed effect, weighted by county population, independently for each date of the pandemic.

## Results

### Vulnerability is not equally distributed across the US

Figure 2 shows the overall vulnerability distributed across 72,173 census tracts within 3,142 counties and 51 states (including the District of Columbia). We find that eight out of the top 10 most vulnerable census tracts reside in Florida with five of those tracts being in the city of Miami. However, vulnerability, according to the CCVI, is overwhelmingly concentrated in the Southern region of the United States where 74% of the Southern population live in a county within the top 2 quintiles of the index (CCVI >= 0.6). We find that seven of the 10 most vulnerable counties are in North Carolina and seven out of the top ten most vulnerable states are located within the South. The overall CCVI can be broken down into scores for each of the seven themes. Appendix 4 shows vulnerability maps of each theme which reveal distinct patterns for each dimension of vulnerability, and precisionforcovid.org/ccvi hosts an interactive version of the index. A breakdown of the variance contributed to the index by each theme using correlation analysis and principal components analysis is shown in Appendix 5.

**Figure 2:**
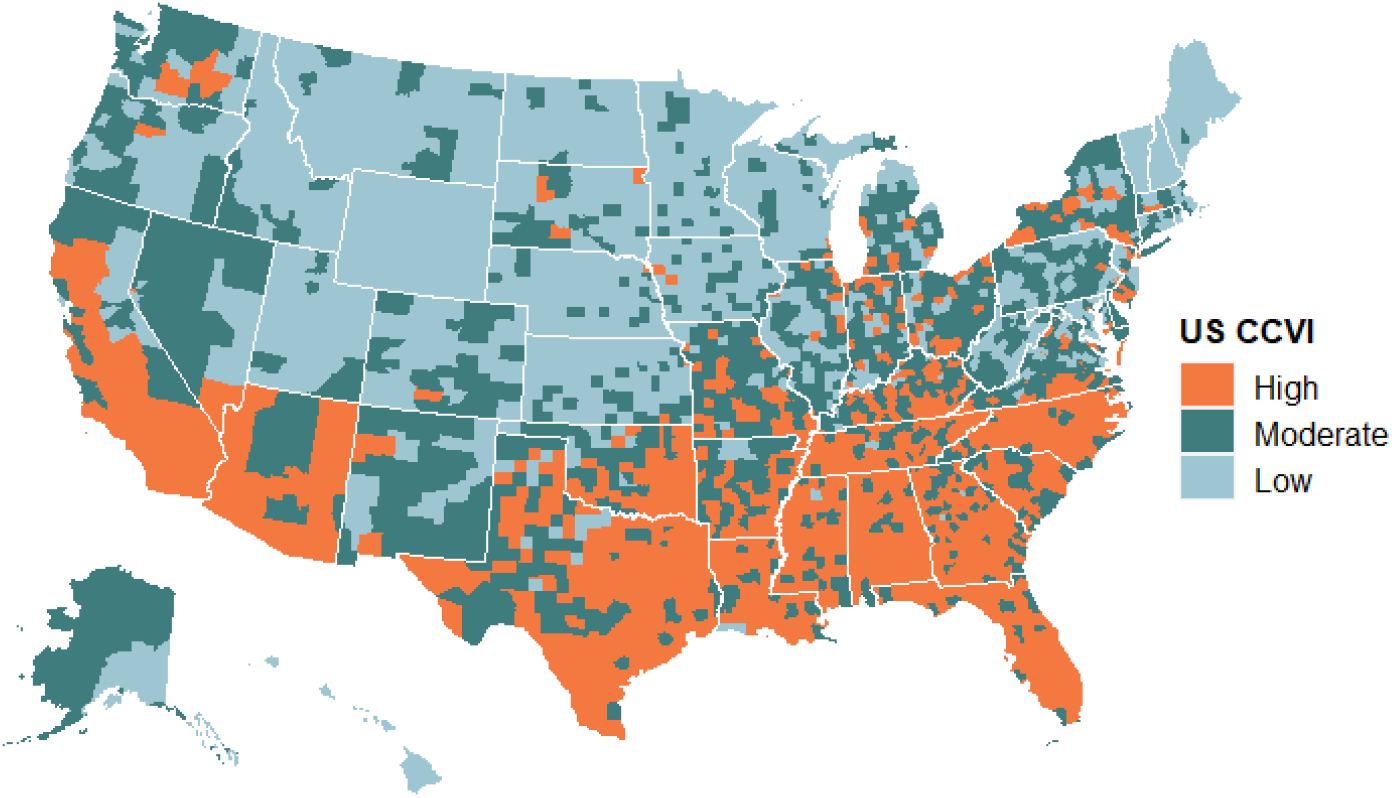
Vulnerability across the US expressed relative to all counties. The CCVI is split into terciles (High, Moderate, Low).

High vulnerability afflicts counties both small and large. The top 10 most populous counties in the US with a CCVI score in the top quintile (i.e. > 0.8, very high vulnerability) have a combined population that equals ~12% of the US. While many vulnerable counties are found in the South, Figure 3 shows other regions, including large cities in the Northeast, Midwest, and the West, hold pockets of vulnerability as well.

**Figure 3.**
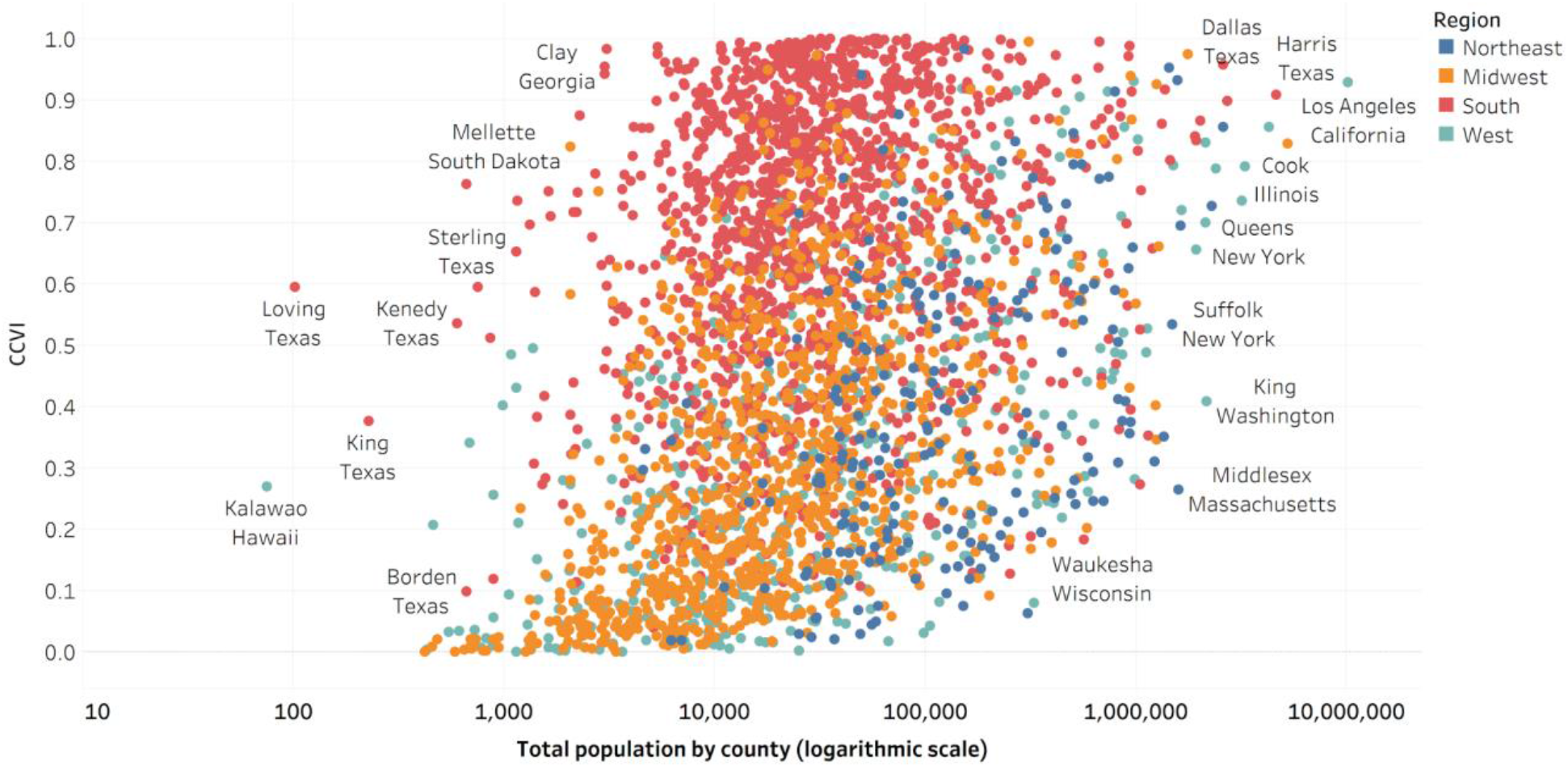
Scatter plot showing every US county as a function of its vulnerability score and its population size. Each dot represents a county. The x-axis is on a logarithmic scale. CCVI: COVID-19 Community Vulnerability Index.

### Vulnerable counties have been harder hit by infections and deaths

As of May 13, 2021, people in vulnerable communities have been 21% more likely to be diagnosed with COVID-19, and 47% more likely to have died, unadjusted for age and comorbidities. These differences were notable during the May and July 2020 peaks, and this inequity has been sustained through almost all of the pandemic, although it decreased briefly during the wave in early winter 2020 as the virus surged in less vulnerable counties as well (Figure 4). As of March 2021, high vulnerability counties had much higher rates of cases and death, demonstrating that vulnerability continues to be a necessary lens through which to monitor national COVID-19 trends. Breaking down vulnerability by seven themes, we find that the impact on different types of vulnerable communities has varied greatly over time (Appendix 6).

**Figure 4.**
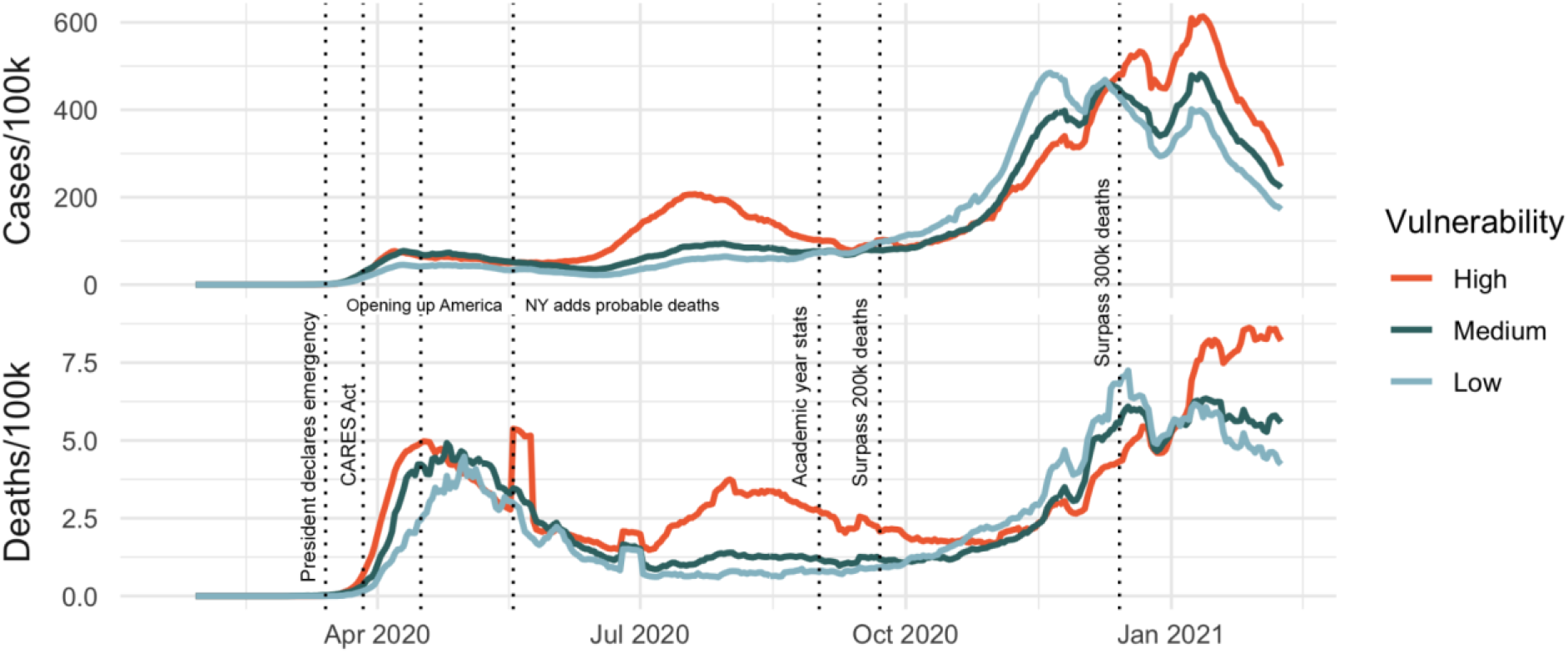
A time series of COVID-19 cases and deaths as a function of vulnerability. Top: New cases per 100k people in the 7 preceding days reported at the county level and disaggregated by vulnerability tercile. Bottom: Deaths per 100k people as a 7-day rolling average (within the past week) reported at the county level and disaggregated by vulnerability tercile. Note the peak around late May was due to the addition of a large number of “probable” COVID-19 deaths.

### Access to test sites is mostly equitable

Examining equity in test site access is crucial to understanding possible bias in reported case data as well as the equity of the COVID-19 response. In urban areas, the CCVI does not have a significant relationship with either test site density (p = 0.57) or distance (p = 0.34). In rural areas, greater vulnerability is associated with increased access (more test sites, p = 0.0002, and shorter travel distances, p = 0.0008), underscoring the progress made in supporting vulnerable rural areas since early gaps in access were noted during summer 2020 (26).

### Rent payments are furthest behind in vulnerable states

Vulnerability has played a key role in risk of various economic outcomes during the COVID-19 pandemic. Decreasing consumer spending and rising unemployment were more dramatically felt by low-wage workers compared to high-wage (2). Due to cascading economic impacts, housing payments are at serious risk of delinquency, with only national and state moratoria stemming mass evictions and foreclosures (27). Across three waves of the U.S. Census Bureau’s Household Pulse Survey between August and December 2020, respondents from states with higher CCVI were more likely to state they were behind on rent (Figure 5). A 10 percentage-point increase in vulnerability is associated with a 0.7 point increase in the percentage of households who are behind on rent payments (p = 0.003). Although levels of delinquency rose across waves, the association with the CCVI does not significantly change over time.

**Figure 5:**
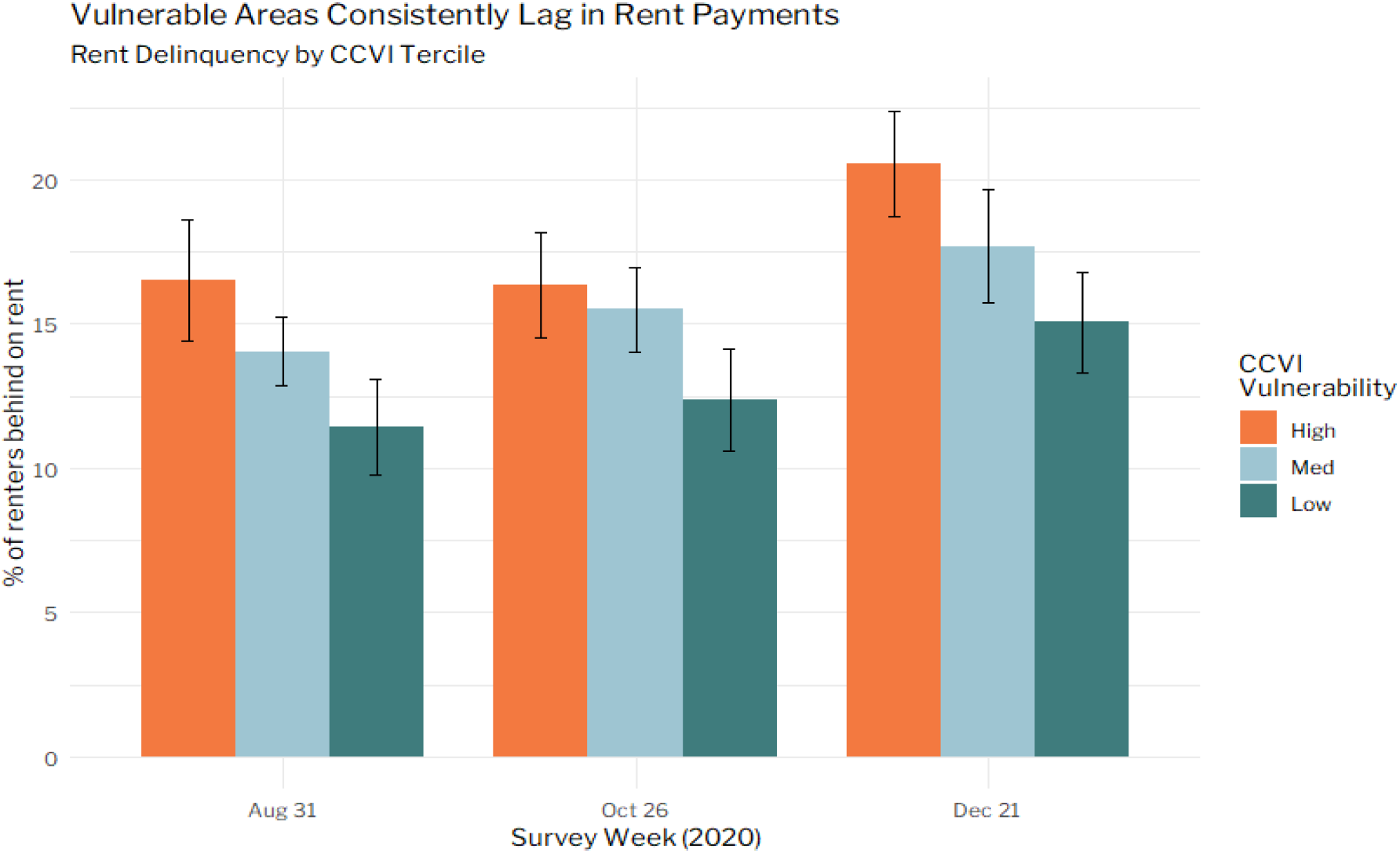
An increasing number of people are falling behind on rent payments, with rates exceeding 20% in highly vulnerable states in December 2020. Error bars represent 95% confidence interval.

### COVID-19 themes in the CCVI are geographically distinct from social vulnerability

Both the CCVI and the CDC’s SVI were designed to help direct resources for communities in need of support during a public health emergency. A population-weighted correlation of SVI and CCVI shows a strong positive relationship between the indices at the county (*r* = .82, *p* < .001) and census tract (*r* = .86, *p* < .001) level (Appendix 7). The SVI and CCVI constituent themes are largely positively correlated with each other, though much less so on the CCVI’s COVID-specific themes, underscoring the importance of considering vulnerabilities beyond social vulnerability alone (Appendix 7).

## Discussion

This paper presents a census tract-level U.S. index capturing multiple drivers of COVID-19 vulnerability. We find that communities identified as vulnerable by this index suffered substantially more COVID-19 cases, higher rates of mortality, and are facing higher rates of rental and mortgage arrears. In contrast, physical test site access is now similar between vulnerable and non-vulnerable populations, after being inequitable earlier in the pandemic. The CCVI builds on the existing Social Vulnerability Index by adding several COVID-19 specific themes that only weakly correlate with social vulnerability.

Using case, death, testing, and economic data, we examined the impact of COVID-19 vulnerability at different points of the pandemic. Inequity in case and death rates is evident across the course of the pandemic, and we have previously reported on inequity in COVID-19 hospitalizations and strain on hospital capacity (28). It was already known that certain county-level factors associate with mortality (29, 30) and social vulnerability is associate with higher mortality early in the pandemic (31), and our results show inequities have persisted throughout most of the first year of the pandemic. Previous work had revealed the disproportionate impact of COVID-19 on low-wage workers (2). We further observe that regions identified as vulnerable by the CCVI also have the highest rates of rent delinquency. In contrast, access to test sites was mostly equitable, unlike earlier in the pandemic (26). This encouraging finding reflects widespread efforts to improve access, such as investment in testing at Historically Black College and Universities (32).

We compared the CCVI against the widely used Social Vulnerability Index, which was developed by the CDC to support disaster management (15). Our intention was to build an index that incorporated elements of social vulnerability known to be critical to virtually any disaster, and add elements specific to COVID-19. We find that these COVID-19-specific themes show markedly different geographic distributions across the U.S. and are only weakly correlated with social vulnerability. Although the overall CCVI and SVI scores are correlated (with roughly two-thirds of variance across counties shared between them), the themes within the CCVI provide the user with far more actionable drivers of vulnerability to mitigate against. The COVID-19-specific themes further showed distinct relationships with mortality over the course of the pandemic, further emphasizing that the additional themes in the CCVI are warranted for this pandemic.

The decision to consider overall vulnerability or domain-level vulnerability using individual themes depends on the specific use case; for example, priority distribution of medical resources might consider healthcare system vulnerability, whereas the establishment of contact-tracing mechanisms might be informed by the concentration of high-risk environments in a given community. The appropriate response to undesirable outcomes varies dramatically based on features of the community, and taking into account the vulnerability profile of a given community allows local governments to target resources efficiently for maximum impact at each phase of the response. Appendix 9 describes some use cases of the CCVI, and lessons learned for constructing and applying an index such as the CCVI for an effective pandemic response.

### Limitations

The CCVI assigns one score to each geographic unit, which contains anywhere between a few thousand to millions of individuals (for a select few counties). However, some indicators are available only at the county or state level, such that each census tract is assigned the same score for that indicator. Whilst this is not always an issue - e.g. health system indicators are not sensible at the census tract level - in some cases this hides substantial heterogeneity for an indicator within a geography. A related point is that every indicator is a summary statistic, hiding large individual differences. There are many counties with enormous inequity in health and social indicators, and the plight of vulnerable populations in such communities is easily lost in a population average.

The index captures underlying vulnerabilities to COVID-19 that change over months or years, and most indicators are based on pre-pandemic data. This means the index does not reflect the pandemic response, such as state-level policies intended to protect the vulnerable, nor does it reflect the day-to-day changes in cases, deaths, mobility, and so forth. This marks a clear difference to many epidemiological models narrowly focused on infections, hospitalization, and mortality (33, 34). However, in our work with government and other partners, the unchanging nature of the index has proven particularly beneficial, as long as the index was appropriately combined with live data. That is, we consider the index a starting point for further analysis, serving as a lens through which to understand (in)equity across surveillance, forecasting, and pandemic response data, none of which our index seeks to replace. A more pragmatic point is that most health departments would not be able to deal operationally with an index updated on a daily or weekly basis.

Finally, the index does not cover U.S. territories as many indicators - including social vulnerability metrics - were not available for these geographies.

## Data Availability

The COVID-19 Community Vulnerability Index and an interactive data explorer are available at the Surgo Venture website. The CDC Social Vulnerability Index is available through the CDC and ATSDR. Confirmed COVID-19 cases and death counts were obtained on March 2, 2021 from the Johns Hopkins University Center for Systems Science and Engineering, which collects and reports COVID-19 data for each US county. COVID-19 testing location data were compiled from GISCorps. Rental arrears at the state level were collected through the U.S. Census Bureau Household Pulse Survey.

https://precisionforcovid.org/ccvi

https://coronavirus.jhu.edu/map.html

https://www.giscorps.org/covid-19-testing-site-locator/

https://www.census.gov/programs-surveys/household-pulse-survey/data.html

https://www.atsdr.cdc.gov/placeandhealth/svi/index.html

## References

1. Hay SI. COVID-19 scenarios for the United States. medRxiv. 2020:2020.07.12.20151191.

2. Chetty R, Friedman JN, Hendren N, Stepner M, Team TOI. How Did COVID-19 and Stabilization Policies Affect Spending and Employment? A New Real-Time Economic Tracker Based on Private Sector Data. National Bureau of Economic Research Working Paper Series. 2020;No. 27431.

3. Czeisler MÉ, Lane RI, Petrosky E, Wiley JF, Christensen A, Njai R, et al. Mental Health, Substance Use, and Suicidal Ideation During the COVID-19 Pandemic - United States, June 24-30, 2020. MMWR Morb Mortal Wkly Rep. 2020;69(32):1049–57.

4. Maringe C, Spicer J, Morris M, Purushotham A, Nolte E, Sullivan R, et al. The impact of the COVID-19 pandemic on cancer deaths due to delays in diagnosis in England, UK: a national, population-based, modelling study. The Lancet Oncology. 2020;21:1023–34.

5. Khazanchi R, Beiter ER, Gondi S, Beckman AL, Bilinski A, Ganguli I. County-Level Association of Social Vulnerability with COVID-19 Cases and Deaths in the USA. Journal of General Internal Medicine. 2020;35(9):2784–7.

6. Mongey S, Pilossoph L, Weinberg A. Which Workers Bear the Burden of Social Distancing Policies? National Bureau of Economic Research Working Paper Series. 2020;No. 27085.

7. Furceri D, Loungani P, Ostry JD, Pizzuto P. Will Covid-19 affect inequality? Evidence from past pandemics. Covid Economics. 2020;12(1):138–57.

8. Qualls N, Levitt A, Kanade N, Wright-Jegede N, Dopson S, Biggerstaff M, et al. Community Mitigation Guidelines to Prevent Pandemic Influenza - United States, 2017. MMWR Recomm Rep. 2017;66(1):1–34.

9. Desmond-Hellmann S. Progress lies in precision. Science. 2016;353(6301):731-.

10. Chandrasekhar R, Sloan C, Mitchel E, Ndi D, Alden N, Thomas A, et al. Social determinants of influenza hospitalization in the United States. Influenza Other Respir Viruses. 2017;11(6):479–88.

11. Lowcock EC, Rosella LC, Foisy J, McGeer A, Crowcroft N. The social determinants of health and pandemic H1N1 2009 influenza severity. Am J Public Health. 2012;102(8):e51–e8.

12. Roberts JD, Tehrani SO. Environments, Behaviors, and Inequalities: Reflecting on the Impacts of the Influenza and Coronavirus Pandemics in the United States. International Journal of Environmental Research and Public Health. 2020;17(12):4484.

13. Covid TC, Team R. Severe Outcomes Among Patients with Coronavirus Disease 2019 (COVID-19)-United States, February 12-March 16, 2020. MMWR Morb Mortal Wkly Rep. 2020;69(12):343–6.

14. America’s Health Rankings | AHR: United Health Foundation; [Available from: https://www.americashealthrankings.org/.

15. Flanagan BE, Gregory EW, Hallisey EJ, Heitgerd JL, Lewis B. A social vulnerability index for disaster management. Journal of homeland security and emergency management. 2011;8(1).

16. NHSPI | National Health Security Preparedness Index: Robert Wood Johnson Foundation; [Available from: https://nhspi.org/.

17. Remington PL, Catlin BB, Gennuso KP. The county health rankings: rationale and methods. Population health metrics. 2015;13(1):11.

18. Burse N, Thompson E, Monger M. The Role of Public Health in COVID-19 Emergency Response Efforts From a Rural Health Perspective. Prev Chronic Dis. 2020;17.

19. Centers for Disease Control and Prevention. COVID-19 Secondary data and statistics 2020 [updated May 6. Available from: https://www.cdc.gov/library/researchguides/2019novelcoronavirus/datastatistics.html.

20. Flanagan BE, Hallisey EJ, Adams E, Lavery A. Measuring community vulnerability to natural and anthropogenic hazards: the Centers for Disease Control and Prevention’s Social Vulnerability Index. Journal of environmental health. 2018;80(10):34.

21. Dong E, Du H, Gardner L. An interactive web-based dashboard to track COVID-19 in real time. The Lancet infectious diseases. 2020;20(5):533–4.

22. Corps G. COVID-19 Testing and Vaccination Sites Data Creation Project. In: ArcGIS, editor. 2021.

23. Bureau UC. Household Pulse Survey: Measuring Social and Economic Impacts during the Coronavirus Pandemic. 2021.

24. Lumley T. Analysis of complex survey samples. J Stat Softw. 2004;9(1):1–19.

25. Lê S, Josse J, Husson F. FactoMineR: an R package for multivariate analysis. Journal of statistical software. 2008;25(1):1–18.

26. Surgo Foundation. Medium [Internet]2020. Available from: https://medium.com/@surgofoundation/millions-of-vulnerable-rural-americans-live-in-covid-19-testing-deserts-e11a64961175.

27. Parrott J, Zandi M. Averting an Eviction Crisis.: Urban Institute; 2021.

28. Baer J, Campigotto C, Coome L, Dibner-Dunlap A, Sgaier SK, Smittenaar P, et al. Vulnerable Communities and COVID-19: The Damage Done, and the Way Forward. Surgo Ventures; 2021.

29. Desmet K, Wacziarg R. Understanding Spatial Variation in COVID-19 across the United States. National Bureau of Economic Research; 2020. Report No.: 0898-2937.

30. Knittel CR, Ozaltun B. What Does and Does Not Correlate with COVID-19 Death Rates. National Bureau of Economic Research Working Paper Series. 2020;No. 27391.

31. Islam N, Lacey B, Shabnam S, Erzurumluoglu A, Dambha-Miller H, Chowell G, et al. Social inequality and the syndemic of chronic disease and COVID-19: county-level analysis in the USA. Journal of Epidemiology Community Health. 2021;75:496–500.

32. Golston A. Why Black colleges and universities are America’s newest–and most critical–diagnostic testing hubs: Bill & Melinda Gates Foundation; 2020 [

33. Zhou Y, Wang L, Zhang L, Shi L, Yang K, He J, et al. A spatiotemporal epidemiological prediction model to inform county-level COVID-19 risk in the United States. Special Issue 1-COVID-19: Unprecedented Challenges and Chances. 2020.

34. Covid I, Murray CJ. Forecasting COVID-19 impact on hospital bed-days, ICU-days, ventilator-days and deaths by US state in the next 4 months. MedRxiv. 2020.

