## Supplemental Material for "A COVID-19 Community Vulnerability Index to drive precision policy in the US"

### Appendix

#### Appendix 1: CCVI Components and Data Sources

*Table 1. COVID-19 Community Vulnerability Index (CCVI) Data sources for indicators and subthemes grouped into seven primary themes.*

| Theme | | Variable | Indicator(s) | Geo Precision | Source |
|---|---|---|---|---|---|
| **1** | **Socioeconomic Status** | Below Poverty | Persons below poverty estimate | Census Tract | CDC, American Community Survey (ACS), 2014-2018 5-Year Estimates |
| | | Unemployed | Civilian (age 16+) unemployed estimate | | |
| | | Income | Per capita income estimate | | |
| | | No High School Diploma | Persons with no high school diploma (age 25+) estimate | | |
| | | Uninsured | Percent of population uninsured | | |
| **2** | **Minority Status & Language** | Minority | Minority (all persons except white, non-Hispanic) estimate | | |
| | | Speaks English “Less than Well” | Persons (age 5+) who speak English "less than well" estimate | | |
| **3** | **Housing type, Transportation, Household Composition & Disability** | Multi-Unit Structures | Housing with structures with 10 or more units estimate | | |
| | | Crowding | Households with more people than rooms estimate | | |
| | | No Vehicle | Households with no vehicle available estimate | | |
| | | Group Quarters | Persons in institutionalized group quarters estimate | | |
| | | Aged 17 or Younger | Persons aged 17 and younger estimate | | |
| | | Single-Parent Households | Single parent households with children under 18 estimate | | |
| | | Access to Indoor Plumbing | Households without access to indoor plumbing | | |
| | | Mobile Homes | Mobile homes estimate | | |
| | | Older than Age 5 with a Disability | Civilian noninstitutionalized population with a disability estimate | | |

| | | | | | |
|---|---|---|---|---|---|
| 4 | **Epidemiological Factors** | Cardiovascular Conditions | Estimated percent of adults diagnosed with high cholesterol | Census Tract (2017) | PolicyMap: CDC, Behavioral Risk Factor Surveillance System (BRFSS), 2017-2018 |
| | | | Estimated percent of adults diagnosed with a stroke | Census Tract (2018) | |
| | | | Estimated percent of adults ever diagnosed with heart disease | | |
| | | Respiratory Conditions | Estimated percent of adults diagnosed with chronic obstructive pulmonary disease, emphysema, or chronic bronchitis | Census Tract (2018) | |
| | | | Estimated percent of adults reporting to smoke cigarettes | | |
| | | Immuno-compromised | Annual cancer incidence per 100,000 persons | County | PolicyMap: CDC, National Cancer Institute (NCI), 2011-2015 |
| | | | Rate of persons living with an HIV diagnosis per 100,000 people | | PolicyMap: CDC, National Center for HIV, STD and TB Prevention (NCHSTP), Division of STD/HIV Prevention, 2016 |
| | | Obesity | Estimated percent of adults reporting to be obese (a body mass index of 30 or greater) | Census Tract (2018) | PolicyMap: CDC, Behavioral Risk Factor Surveillance System (BRFSS), 2018 |
| | | Diabetes | Estimated percent of adults ever diagnosed with diabetes | Census Tract (2018) | |
| | | Aged 65 or Older | Persons aged 65 and older estimate | Census Tract | CDC, American Community Survey (ACS), 2014-2018 5-Year Estimates |
| 5 | **Healthcare System Factors** | Health System Capacity | Intensive Care Unit (ICU) Beds per 100,000 | County | Kaiser Health News, Centers for Medicare, & Medicaid Services (CMS), 2018-2019 |
| | | | Hospital Beds per 100,000 | | Definitive Healthcare, 2020 |
| | | | Epidemiologists per 100,000 | State | U.S Bureau of Labor Statistics (BLS), Occupational Employment and Wages, May 2018 |
| | | Health System Strength | Agency for Healthcare Research and Quality - Prevention Quality Indicator Overall Composite (PQI): admission rates for preventable conditions (via good outpatient care) adjusted per population | County | Centers for Medicare, & Medicaid Services (CMS), Mapping Medicare Disparities (MMD) Tool, 2017 |

| | | | | | |
|---|---|---|---|---|---|
| | | | Health Spending per Capita | State | Centers for Medicare, & Medicaid Services (CMS), Health Expenditures by State of Residence, 2014 |
| | | | Aggregate cost of medical care | Census Tract | PolicyMap & Quantitative Innovations, 2017 |
| | | Healthcare Accessibility | Percent of population with a Primary Care Physician | Census Tract | PolicyMap & CDC BRFSS, 2018 |
| | | Health System Preparedness | Total Public Health Emergency Preparedness (PHEP) Funding Per Capita | State | CDC, Center for Preparedness and Response, 2019 |
| | | | Health Labs per 100,000 | County | Association of Public Health Laboratories |
| **6** | **High Risk Environments** | Percentage of population working or living in environments with high infection risk | Long-term care (nursing homes, assisted living, and care homes) residents per 100,000 | Census tract | ArcGIS/Dept of Homeland Security |
| | | | Prisons population per 100,000 | County | Vera institute for Justice, 2016 |
| | | | Percentage of population employed in high-risk industry | County | BLS QCEW 2020 |
| **7** | **Population Density** | Population Density | Estimated total number of people per unit area (sq. miles) | Census Tract | CDC Social Vulnerability Index |

### Appendix 2: Logic Model

A literature search was conducted up to August 10, 2020 using PubMed and several collated COVID-19 literature resources (1); (2); (3) to identify drivers of vulnerability to pandemic health, economic, and social impacts. Combinations of terms were used including but not limited to: pandemic; risk factors; vulnerability; COVID-19; coronavirus; mortality; severity; transmission; epidemiology; social determinants; chronic conditions; healthcare capacity. Publications were prioritized by type of study, quality, and source, including systematic reviews and meta-analyses, robust primary research, and guidance from governing bodies. This evidence helped formulate the rationale for including each of the 6 themes in the US CCVI as described in the following sections.

#### Socioeconomic Status

During a pandemic, communities with lower socioeconomic status may face additional barriers to recovery with limited access to financial and healthcare resources (4-6) and can experience adverse psychological outcomes due to difficulties coping (7-9). A lower educational level can limit the accessibility to and understanding of recovery information and the likelihood of preventative action (4, 5). During the COVID-19 pandemic, less educated workers have disproportionately experienced poor labor market outcomes (10). Low income individuals are less likely to increase social distancing, hand washing or masking wearing behaviors; these preventative measures may be less practical due to a higher likelihood of working in occupations not conducive to teleworking or experiencing job or income loss (11). Job loss can exacerbate unemployment rates resulting in a large uninsured population, which can lead to decreased care-seeking behavior and added response complexity (5, 12, 13). Higher rates of COVID-19 cases and deaths have been reported in socioeconomically disadvantaged communities with high poverty, unemployment, and lower income and education levels (14-18).

#### Minority Status and Language

Minority populations are more likely to live in crowded settings, work in essential occupations, have lower socioeconomic status, and have a higher prevalence of comorbidities with less access to quality healthcare -- all of which can limit preventative behaviors, including social distancing and care-seeking, and lead to increased spread and greater morbidity and mortality (4, 6, 19). These communities often have less access to recovery resources and may face communication challenges with a greater proportion of non-English speaking households, which complicates sharing of pandemic information and further prohibits preventive action to reduce spread (4, 6). Minority and non-english speaking communities are also at greater risk of adverse psychological outcomes during a pandemic (8, 9). As a result, COVID-19 has disproportionately impacted minority and non-English speaking populations with a greater number of cases, hospitalizations, and deaths (18, 20-24). Minorities have also disproportionately experienced adverse labor market outcomes with greater economic loss and higher unemployment rates and faced food insecurity and difficulties paying rent (10, 25, 26).

#### Housing Type, Transportation, Household Composition & Disability

Individuals living in multi-unit structures, mobile homes, and group quarters are more likely to experience crowding and risk of disease spread (5). Institutionalized populations living in group quarters may also have inadequate water and sanitation conditions, often have less access to recovery resources, and are more likely to experience adverse psychological outcomes (4, 9, 27); COVID-19 cases have spread faster in these settings with both case and death rates higher than US population rates (27, 28). Similarly, COVID-19 case rates have been higher in crowded communities and in those with low household vehicle ownership, which limits ability to social distance and access healthcare services (16, 18, 29-31). Areas with household crowding have also faced more deaths with less benefit from social distancing behavior (14, 32). Both housing type and transportation factors have predicted county-level COVID-19 cases in the US (33).

The elderly and disabled are more likely to require health, financial, and mobility-related support and face greater risk of adverse psychological outcomes during a pandemic (4, 5, 9). Similarly, single-parent households have to take on greater responsibility with less resources and time and limited finances adding difficulty to pandemic coping and recovery (4-6) ; (34). In the context of COVID-19, the elderly are at higher risk of COVID-19 severity, including hospitalization and death (35-38). The disabled are more likely to have underlying conditions, difficulty social distancing, and disproportionate access to care and have a greater proportion of COVID-19 cases in younger populations with higher case fatality rates (39, 40). In previous pandemics, younger children have experienced greater severity and mortality (41, 42). While most COVID-19 cases are asymptomatic and the hospitalization rate is low in young children, one in three hospitalized cases are admitted to the ICU, similar to adult rates (43). Children are recognized as a vulnerable population due to limited knowledge and independence for prevention and coping (5, 42).

#### Epidemiological Factors

Respiratory virus co-infection and chronic conditions including hypertension, cardiovascular disease, obesity, chronic lung disease, and diabetes have been associated with COVID-19 hospitalization, severity, and mortality (35, 44, 45)   (37, 46, 47). Cancer, HIV, and other immunosuppressive or immunodeficient conditions often occur with other COVID-19 risk factors, such as chronic comorbidities, and may increase COVID-19 severity, intensive care hospitalization, and risk of death (48-50); (36). Population density can accelerate COVID-19 transmission and has been associated with greater cases and deaths (51-53).

#### Health System Factors

During pandemics, health systems can be stretched to capacity due to rapid spread, further exacerbated by long incubation and treatment windows (54). Areas with low hospital bed density will face large resourcing gaps in accommodating COVID-19 patients, especially if cases surge or if non-COVID-19 bed occupancy is not reduced (55). Areas with less providers and hospitals have faced more COVID-19 cases and deaths, potentially driven by delays due to limited resources and may face lengthened recovery (4, 56). Capacity is needed for pandemic surveillance and mitigation, of which epidemiologists play an essential role (57). A strong health system is needed to effectively respond to COVID-19, including a high quality of care, which is often difficult to measure but strained with overworked providers in a pandemic setting (58, 59). Preparedness for outbreaks requires funding, public health laboratories, and emergency services to readily and rapidly respond with testing and provision of care for COVID-19 patients (60).

#### High Risk Environments

Differences in living and workplace conditions can put sub-groups of the population at increased risk of contracting COVID-19. These “high risk” conditions - proximity to and interaction with other people, and exposure to diseases - and their geographical distribution can make counties more or less vulnerable to COVID-19. Also, current estimates show that nursing home residents account for ~38% of all COVID-related deaths while only accounting for 5% of the cases (61). This disproportionate ratio in mortality to case load prompts a drastic response and inclusion in a COVID-specific vulnerability index.

Workplace crowding and high exposure environments can also make communities more vulnerable to the effects of COVID-19. For instance, rural counties were particularly affected by outbreaks due to their employment dependence on industries with high-risk work conditions like the meatpacking industry (62). Around 50 counties in the US employ over 20% of their workforce in animal slaughtering and processing. The CCVI captures this community reliance on high risk employment by considering industries where workers are in enclosed spaces for extended time, in proximity of or interacting with others, and/or exposed to diseases. The selection of “high risk” industries was informed by O*Net’s database on workplace conditions by occupation (63).

Theme 6 - high risk environments and jobs - includes indicators for sub-groups of the population whose living or work environments puts them at high risk of contracting the virus:

1. Nursing home and assisted living residents
2. Prison population
3. Workers in high-risk industries. Indicator: Percentage of the county’s population employed in a high-risk industry, where high risk industries were selected based on the frequency and duration of contacts with other people in the workplace. These include meat and poultry processing, manufacturing and passenger ground transportation (see table below).
    a. Filtered by % of the population of the county that a specific industry concentrates.
        i. A community (i.e. county) that is dominated by one type of employer (manufacturing/production/etc.)

High-risk industries included

| Industry NAICS Code | Short name |
|---|---|
| 311 | Food_mfg |
| 312 | Beverage_tobacco_mfg |
| 313 | Textile_mills |
| 314 | Textile_product_mills |
| 315 | Apparel_mfg |
| 316 | Leather_and_product_mfg |
| 321 | Wood_product_mfg |
| 322 | Paper_mfg |
| 323 | Printing_and_support_activities |
| 324 | Petroleum_coal_mfg |
| 325 | Chemical_mfg |
| 326 | Plastics_rubber_mfg |

| | |
|---|---|
| 327 | Nonmetallic_mineral_product_mfg |
| 331 | Primary_metal_mfg |
| 332 | Fabricated_metal_product_mfg |
| 333 | Machinery_mfg |
| 334 | Computer_and_electronic_product_mfg |
| 335 | Electrical_equipment_and_appliance_mfg |
| 336 | Transportation_equipment_mfg |
| 337 | Furniture_and_related_product_mfg |
| 339 | Miscellaneous_mfg |
| 485 | Transit_ground_passenger_transport |

#### Population Density

Population density has been shown to be a good predictor of dynamic health outcomes influenced by COVID-19 throughout the evolution of the pandemic specifically in the US. Population density not only increases the total number of people in a defined area, but it also presents the opportunity for more interactions among the population because there is more connectivity among the population. The US has had much more lenient restrictions regarding mobility throughout the pandemic compared to other countries around the world, which could explain why population density is a bigger factor in health outcomes in the US specifically. Other countries with more stringent measures regarding decreased mobility among its residents may be able to mitigate the impact population density could have on the transmission of the virus. However, in the US, these measures were not as strict, thereby creating continued opportunities for the transmission of the virus in denser communities.

There have been studies suggesting that population density is not the sole factor contributing to the adverse COVID-related impacts in cities (64). These studies suggest that other factors such as socioeconomic status, crowding, and total metropolitan size are confounding factors that should be considered. The CCVI also accounts for these and more confounding factors, and together with population density can create a holistic view into vulnerability throughout the pandemic.

We also ran regression analyses with individual indicators against COVID-specific outcomes, and population density consistently was one of the top predictors of adverse health-outcomes justifying its inclusion in the index.  While population density may not be the “exact” or “only” indicator leading to adverse COVID outcomes, it certainly warrants inclusion in the CCVI index.

### Appendix 3: Aggregation of indicators and themes

We followed the CDC SVI's methodology for transforming indicators onto a common scale and aggregating them into themes and an overall score through a series of percentile rank and aggregation steps (5). The census tract CCVI was constructed from census tract indicators, and the county CCVI from county indicators, where applicable. Each indicator was percentile ranked across counties and summed within its theme. This procedure was repeated on theme scores to arrive at an overall CCVI score. This method implicitly weights indicators inversely proportional to the number of other indicators in the theme. The CCVI is a combination of hierarchical and deductive configurations (65). The hierarchical configuration involves conceptually organizing indicators into themes that define various dimensions of vulnerability. However, it is functionally a deductive configuration since the individual indicators are linearly combined and all explicit weighting is ignored.

We questioned if individual factors linked with COVID-19 outcomes should be explicitly upweighted, and whether COVID-19-specific Themes 4, 5, 6, and 7 should contribute more to overall vulnerability. Weighting was evaluated by examining the sensitivity of the index to the weighting structure (66). We performed basic sensitivity analyses to estimate the impact of upweighting COVID-specific indicators within Theme 4, 5, 6 and 7 on their predictive power against a range of COVID-specific outcomes spanning health, economic, and social dimensions. Results showed high correlations between such versions of the index while predictive capabilities for certain economic and social outcomes suffered. Others have similarly found that more advanced approaches to weighting often lead to a similar end result (67, 68). We therefore elected not to use any weighting to construct the CCVI and instead encourage users to use the theme-based structure as well as raw indicators depending on the context of the application.

### Appendix 4: Vulnerability maps by theme

#### Socioeconomic Status

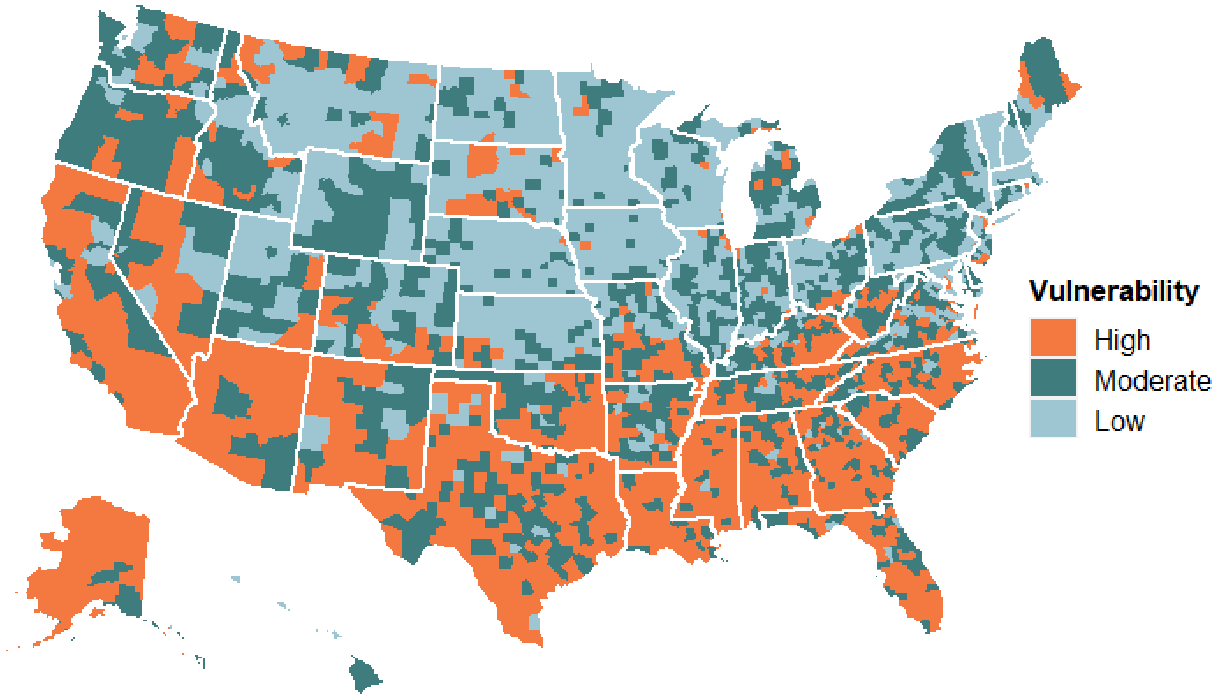


#### Minority Status & Language

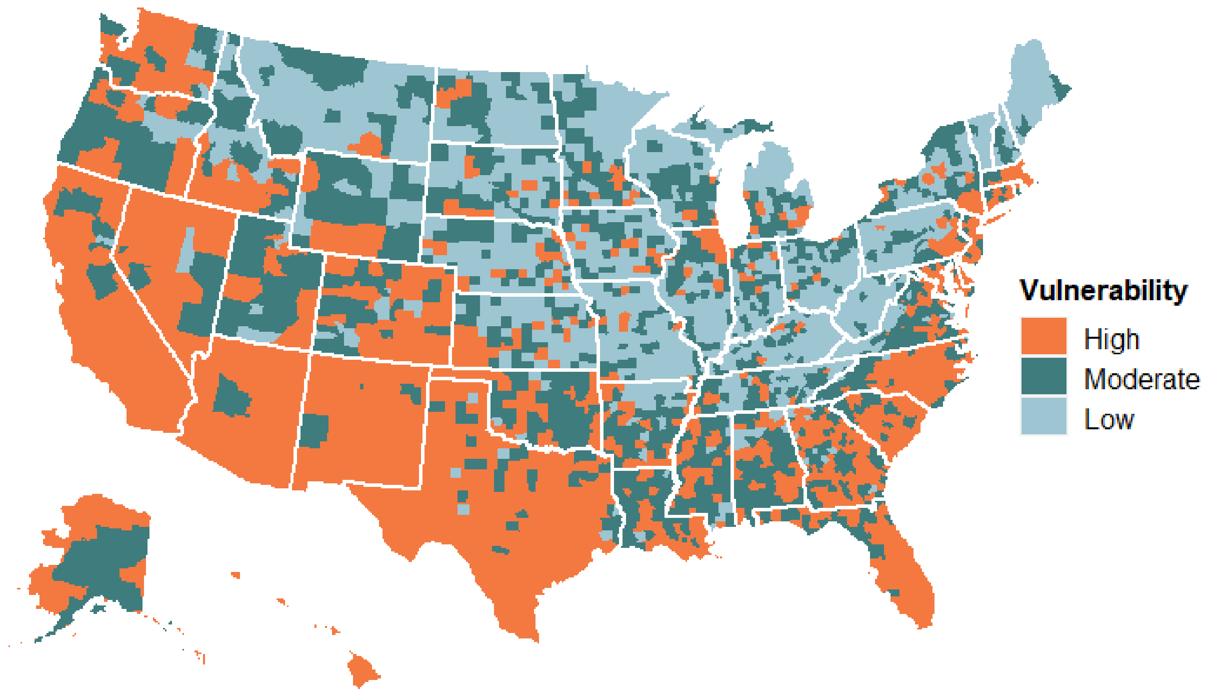

Household Composition & Housing Type

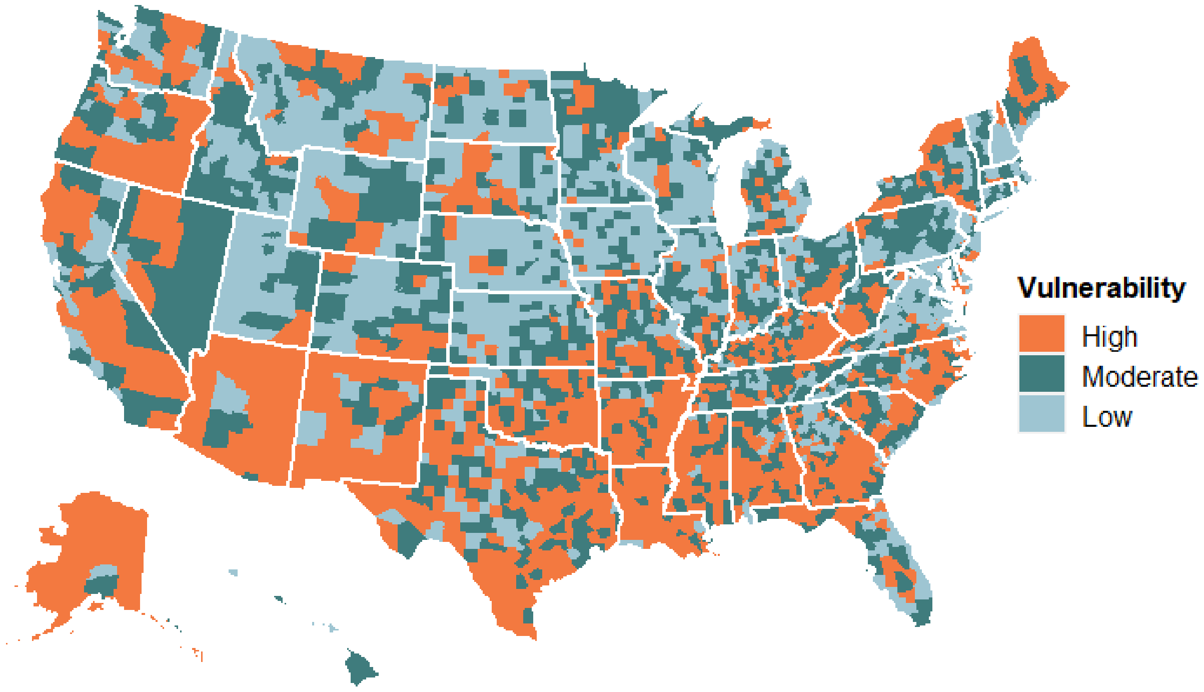


Epidemiological Factors

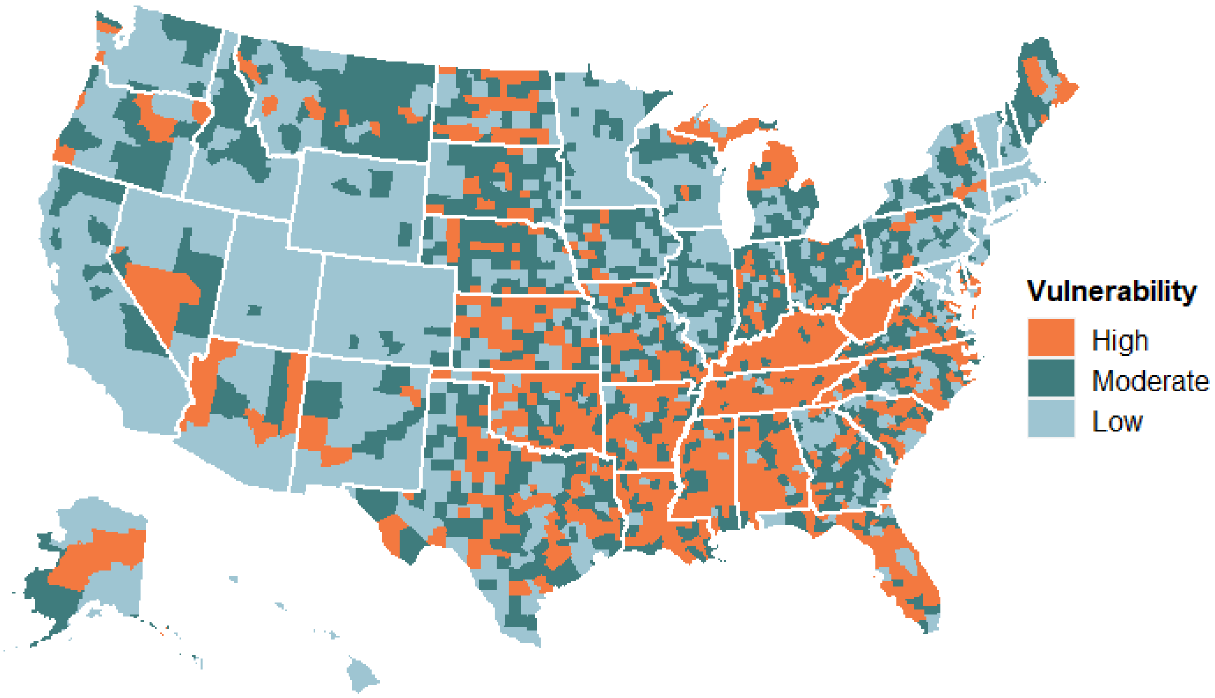

#### Healthcare System Factors

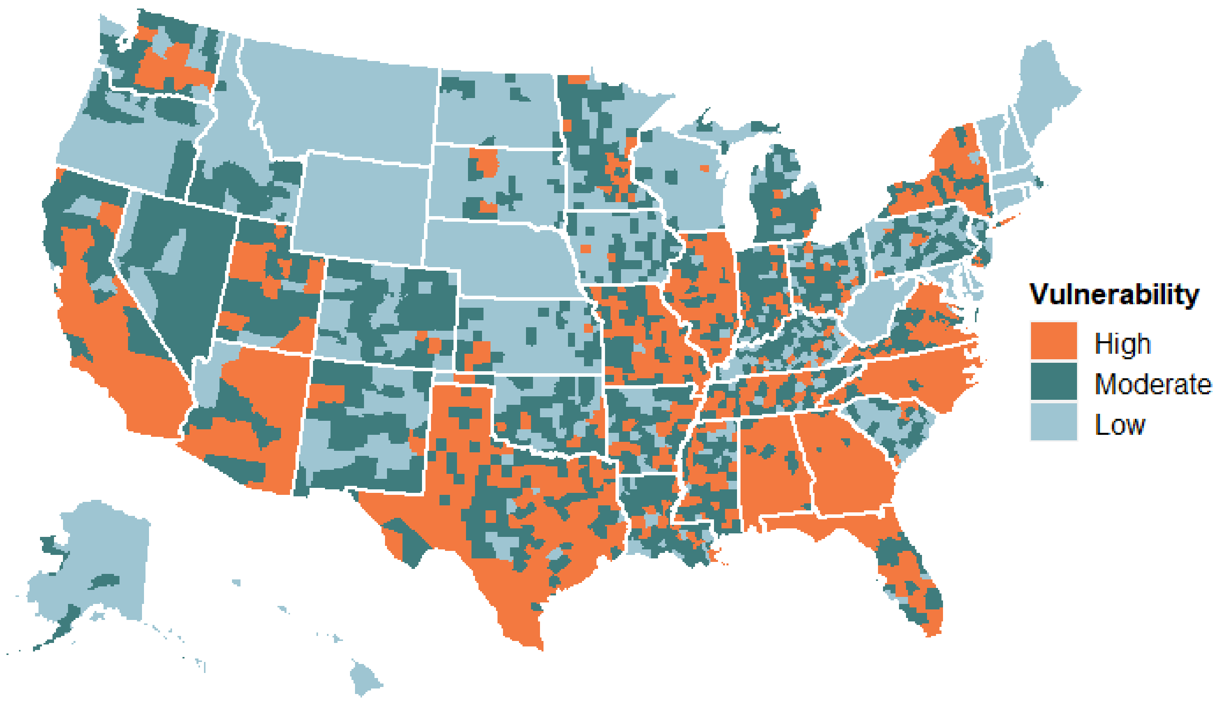


#### High Risk Environments

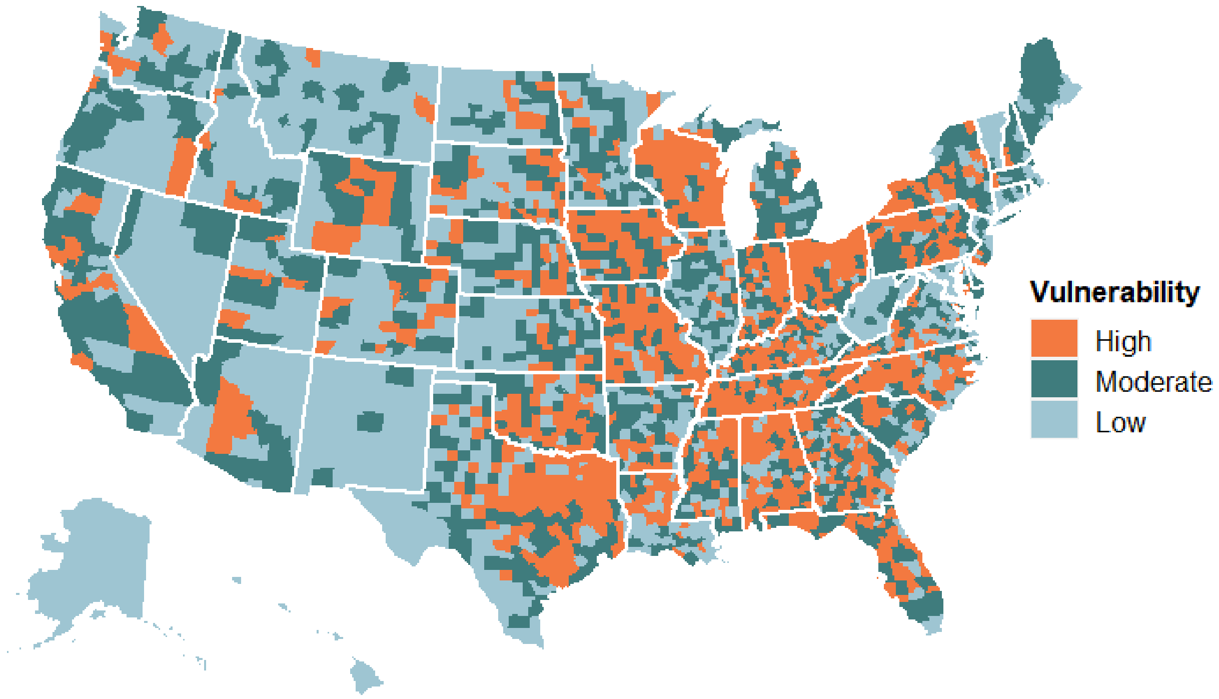

Population Density

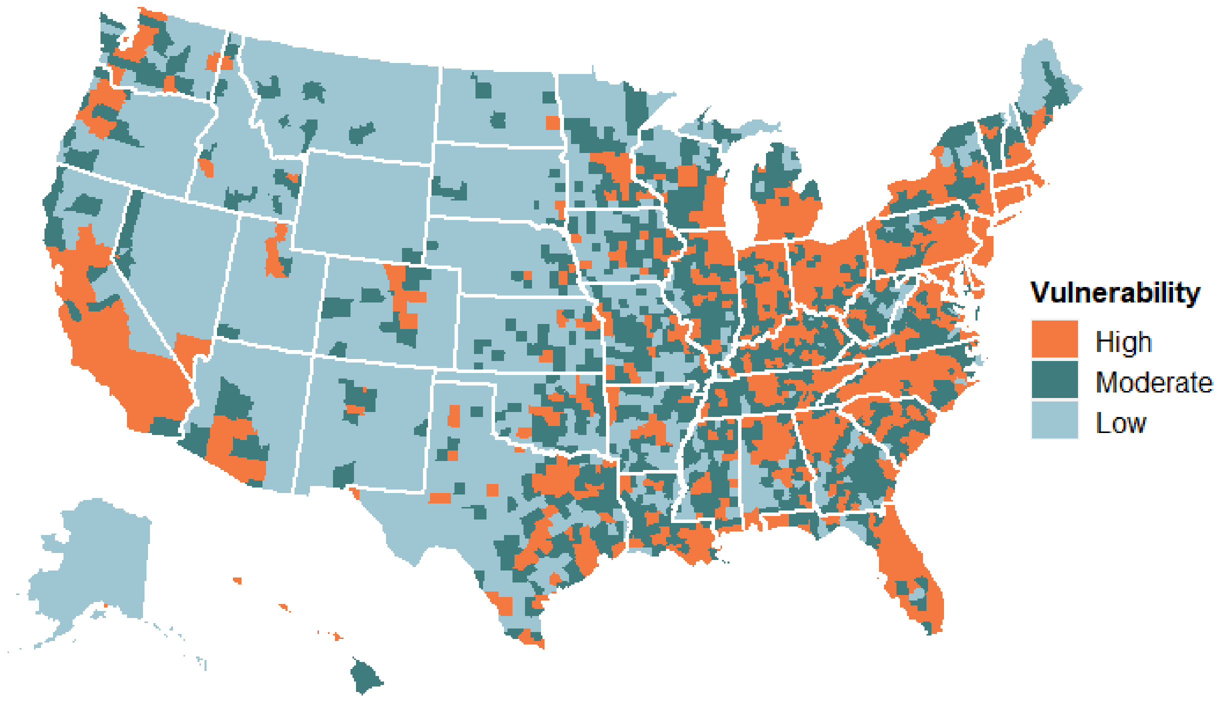


### Appendix 5: Themes contribute distinct variance to the index

We observe that certain types of vulnerabilities tend to co-exist across geographies. The social determinants are mostly positively correlated: socioeconomic vulnerability is positively correlated with minority status and language ($r = 0.50$), and with housing type, transportation, household composition and disability ($r = 0.66$; Figure A). A principal component analysis (PCA) on the themes revealed two major components explaining 60% of the variance (Figure B). The primary component explaining 36% of the variance shows each of the three types of vulnerability from the CDC's SVI, as well as population density and healthcare vulnerability, tend to co-occur. The second component explaining 24% of the variance reflects more rural regions with high vulnerability on all factors except healthcare and minority status. High-risk environments is only weakly associated with both components, suggesting it provides an aspect of vulnerability not readily captured in other themes.

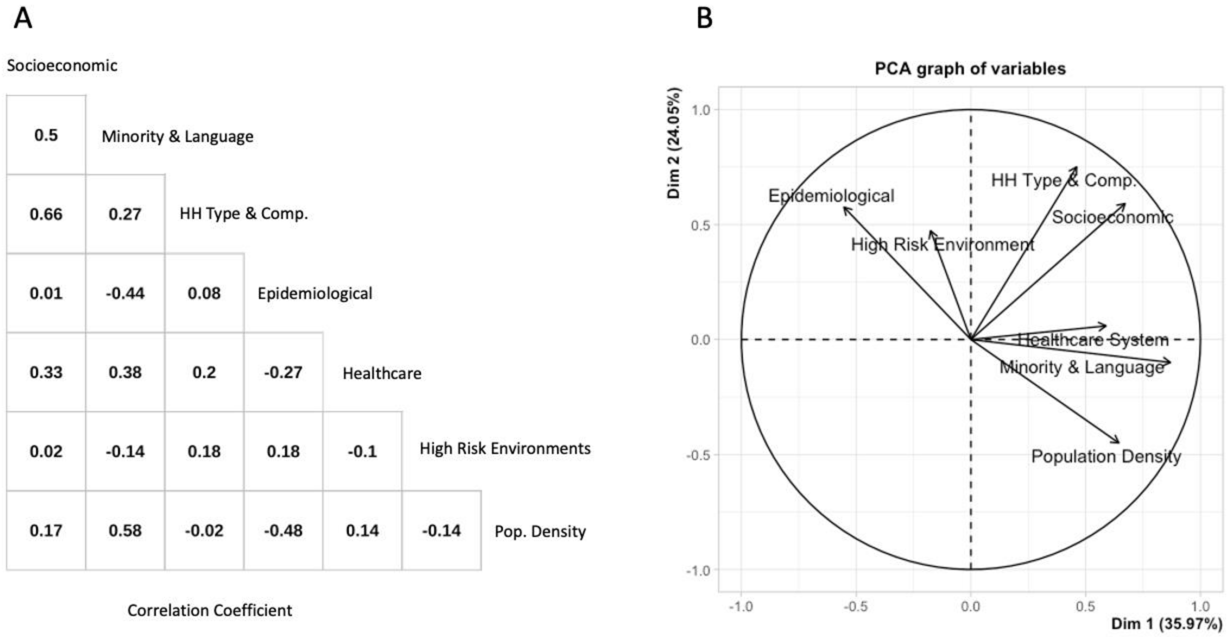


*Correlation and variance decomposition of vulnerability scores relative to other census tracts within the US. A: Correlogram using Pearson correlations. B: Biplot for the first two components in a principal component analysis. The percentages reflect the amount of variance in the index explained by the components. The vectors reflect the loadings of each theme onto the first two principal components.*

### Appendix 6: Communities with different vulnerabilities were hit hardest at different times

As the pandemic swept across the country, different types of communities were impacted in turn. We estimated the relationship between each vulnerability theme with mortality on each day of the pandemic (Figure). In the initial wave, deaths primarily occurred in communities with social vulnerability (high minority representation and/or poor socioeconomic status), poor healthcare systems, and high population density (April-June 2020, positive associations in Figure). In contrast, counties with relatively many in high-risk environments were initially spared (notwithstanding the known impact on nursing homes and other industrial settings). During the Summer months of 2020, relationships with vulnerability were relatively weak though still positive. Then in October, as the winter wave started taking off, a different set of regions were hardest-hit: those with epidemiological risk, housing/transport/disability vulnerability, and high-risk environments. Furthermore, deaths were more likely in rural (i.e. low population density) areas, and those with smaller minority populations. Overall, this shows how there is no constant relationship between specific types of vulnerability and mortality, but rather a pattern that changes over time and by vulnerability type.

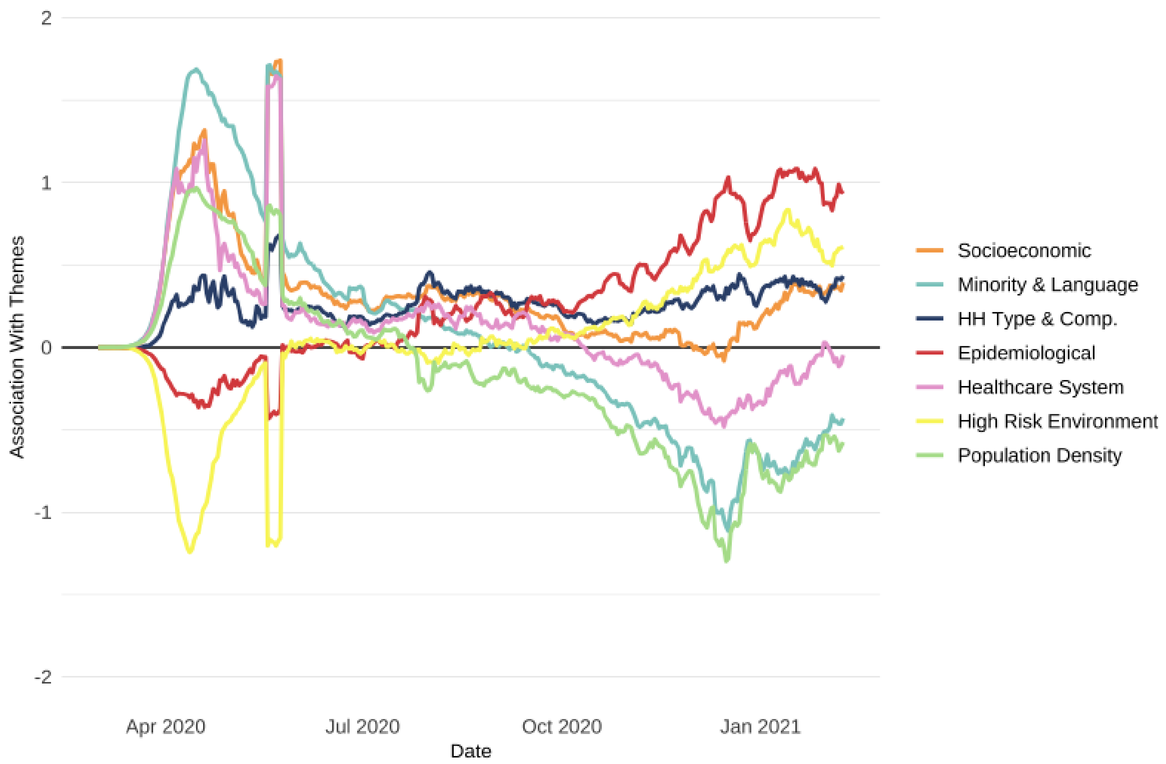


*Population-weighted bivariate regressions of the seven CCVI themes against death rates per 100,000 (7-day average) at the county level with state as a fixed effect, estimated independently for each day in the pandemic. Over the course of the pandemic, different drivers of vulnerability were most strongly related to mortality as the virus swept back and forth across the country. Positive values indicate that more-vulnerable counties experienced higher mortality at that point in time. 95% confidence intervals are omitted for visualization purposes but are approximately +/- .08.*

### Appendix 7: Association between Social Vulnerability Index and COVID-19 Community Vulnerability Index

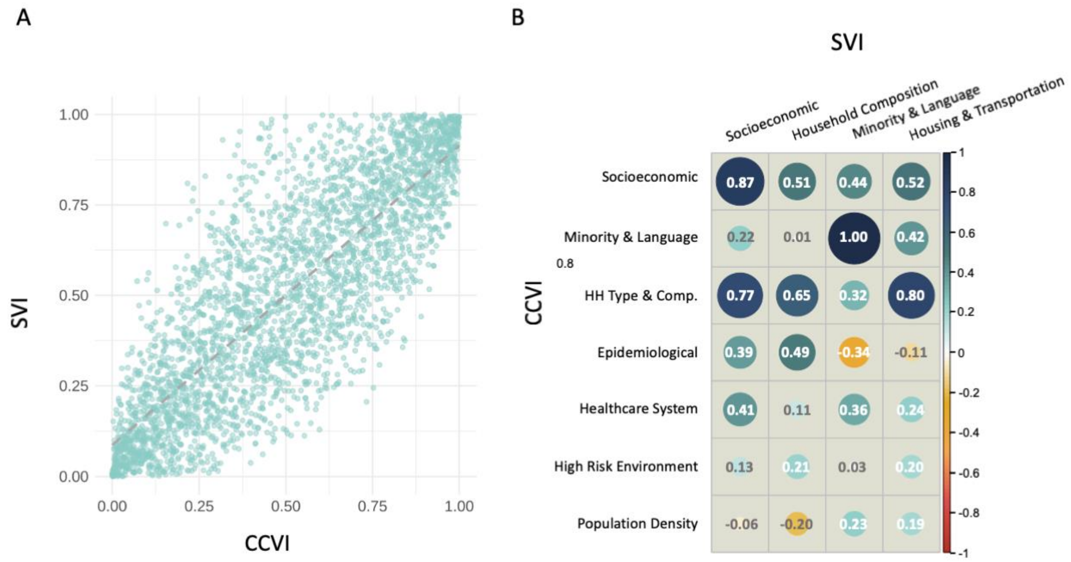


*Comparison of CCVI and SVI. A: Scatterplot of every US county as a function of its CCVI score by its SVI score. Each dot represents a county. Line represents the linear fit between the two scores. B: Correlogram of SVI themes and CCVI themes. Values indicate Pearson correlation coefficients across counties. CCVI: COVID-19 Community Vulnerability Index; SVI: CDC Social Vulnerability Index; HH Type & Comp.: housing type, transportation, household composition, and disability.*

### Appendix 8: Leveraging the CCVI for the COVID-19 Response

To raise CCVI awareness and enable use, we shared CCVI results through several mediums to engage varied audiences. This included website development, press releases, presentations with experts and cross-sector stakeholders, and other media engagements (e.g. journalist interviews and COVID-19 discussion panels). At a national level, the CCVI has been recognized by the CDC (69, 70) and the National Academies of Sciences & Medicine (71) as a valuable resource to guide equitable pandemic response efforts and COVID-19 vaccine allocation. CCVI insights have also informed a bipartisan guideline to efficiently allocate limited resources for the scale up of testing, contact tracing, and supported isolation. In collaboration with Harvard

University, Microsoft, and other private partners, we found that regions with low COVID-19 prevalence but high CCVI vulnerability would gain the most benefit from each test in terms of lives saved per test (72, 73). The index has also informed targeted mitigation strategies to slow spread in vulnerable hotspots and improve educational outcomes in vulnerable areas with poor academic achievement (Nichols, 2020). In addition to the policy applications, the CCVI also generated a number of joint initiatives to develop new tools and analyses locally and globally (74-76).

#### Lessons Learned

Constructing the CCVI in a timely manner proved challenging during a new pandemic with evolving evidence. We share key lessons learned to guide future tool development for precision public health.

**Clearly define the goal for the index.** An early landscape assessment helped to identify key barriers to an effective response, think critically about what value the index could provide, and determine what we were trying to measure and why. A clear definition of the goal - supporting policymakers in a precision COVID-19 response - and outcome measure - resilience to broad COVID-19 impacts - were essential to set parameters for index construction (e.g. rapid tool development) and contextual relevance (e.g. factors linked with health, economic, and social impact).

**Build from a known starting point and engage stakeholders early for buy-in and contextualization.** Using a known, validated starting point (CDC SVI) established credibility and expedited construction. Early consideration of methodological approaches and consultations with field experts helped determine the most appropriate methodology aligned with our goal while understanding limitations for full transparency. While fast, this process was iterative to generate a methodologically sound index in a rapid timeframe and laid foundational partnerships for practical applications of the CCVI.

**Consider the context and end-user needs to design and showcase an actionable, relevant tool.** The CCVI's modular, theme-based approach provides a multi-functional view of vulnerability to help policymakers identify heterogeneity in where and what type of support communities need for a precision public health response. This modular design also allows for modifications, which are expected in a landscape of developing pandemic evidence. Linking the CCVI to dynamic pandemic characteristics helps contextualize COVID-19 impact to continuously guide policymaker decisions.

**Disseminate content strategically through different channels to raise awareness and promote use.** Communication through a variety of mediums over time was necessary to reach stakeholders and demonstrate continued value through practical applications. Communications efforts used clear and concise language to avoid diminishing engagement and information overload. Rather than large, complex analyses, we shared local stories and data, which were more personable and generated the most interest.

**Engage with policy makers directly to stimulate use and build partnerships.** In a rapidly evolving context saturated with a growing evidence base, proactively reaching out to decision-makers to present findings, targeted to their specific objectives and constituencies, allowed us to earn trust and garner valuable feedback. In turn, our policy partners' input was incorporated into new iterations of analysis as well as disseminated through our network of partners in the form of refined applications and use cases for the CCVI.

### References


1. Organization WH. COVID-19 Global literature on coronavirus disease. 2021.
2. Medicine NLo. LitCOVID. 2021.
3. Health NIo. COVID-19 Portfolio. 2021.
4. Cutter SL, Boruff BJ, Shirley WL. Social Vulnerability to Environmental Hazards*. Social Science Quarterly. 2003;84(2):242-61.
5. Flanagan BE, Gregory EW, Hallisey EJ, Heitgerd JL, Lewis B. A social vulnerability index for disaster management. Journal of homeland security and emergency management. 2011;8(1).
6. Hutchins SS, Truman BI, Merlin TL, Redd SC. Protecting vulnerable populations from pandemic influenza in the United States: a strategic imperative. Am J Public Health. 2009;99 Suppl 2(Suppl 2):S243-S8.
7. Cao W, Fang Z, Hou G, Han M, Xu X, Dong J, et al. The psychological impact of the COVID-19 epidemic on college students in China. Psychiatry Research. 2020;287:112934.
8. McGinty EE, Presskreischer R, Han H, Barry CL. Psychological Distress and Loneliness Reported by US Adults in 2018 and April 2020. JAMA. 2020;324(1):93-4.
9. Perrin PC, McCabe OL, Everly GS, Jr., Links JM. Preparing for an influenza pandemic: mental health considerations. Prehospital and disaster medicine. 2009;24(3):223-30.
10. Béland L-P, Brodeur A, Wright T. The short-term economic consequences of Covid-19: exposure to disease, remote work and government response. 2020.
11. Papageorge NW, Zahn MV, Belot M, van den Broek-Altenburg E, Choi S, Jamison JC, et al. Socio-Demographic Factors Associated with Self-Protecting Behavior during the Covid-19 Pandemic. National Bureau of Economic Research Working Paper Series. 2020;No. 27378.
12. Gangopadhyaya A, Garrett AB. Unemployment, Health Insurance, and the COVID-19 Recession. Health Insurance, and the COVID-19 Recession (April 1, 2020). 2020.
13. Woolhandler S, Himmelstein DU. Intersecting U.S. Epidemics: COVID-19 and Lack of Health Insurance. Annals of Internal Medicine. 2020;173(1):63-4.
14. Chen JT, Krieger N. Revealing the Unequal Burden of COVID-19 by Income, Race/Ethnicity, and Household Crowding: US County Versus Zip Code Analyses. Journal of Public Health Management and Practice. 2020.
15. Hawkins D. Social Determinants of COVID-19 in Massachusetts, United States: An Ecological Study. J Prev Med Public Health. 2020;53(4):220-7.
16. Maroko AR, Nash D, Pavilonis BT. COVID-19 and Inequity: a Comparative Spatial Analysis of New York City and Chicago Hot Spots. Journal of Urban Health. 2020;97(4):461-70.
17. Ramírez IJ, Lee J. COVID-19 Emergence and Social and Health Determinants in Colorado: A Rapid Spatial Analysis. International Journal of Environmental Research and Public Health. 2020;17(11):3856.

18. Rozenfeld Y, Beam J, Maier H, Haggerson W, Boudreau K, Carlson J, et al. A model of disparities: risk factors associated with COVID-19 infection. International Journal for Equity in Health. 2020;19(1):126.
19. Tai DBG, Shah A, Doubeni CA, Sia IG, Wieland ML. The Disproportionate Impact of COVID-19 on Racial and Ethnic Minorities in the United States. Clinical Infectious Diseases. 2020.
20. Aldridge R, Lewer D, Katikireddi S, Mathur R, Pathak N, Burns R, et al. Black, Asian and Minority Ethnic groups in England are at increased risk of death from COVID-19: indirect standardisation of NHS mortality data [version 2; peer review: 3 approved]. Wellcome Open Research. 2020;5(88).
21. Khunti K, Singh AK, Pareek M, Hanif W. Is ethnicity linked to incidence or outcomes of covid-19? : British Medical Journal Publishing Group; 2020.
22. Kirby T. Evidence mounts on the disproportionate effect of COVID-19 on ethnic minorities. The Lancet Respiratory Medicine. 2020;8(6):547-8.
23. Moore JT, Ricaldi JN, Rose CE, Fuld J, Parise M, Kang GJ, et al. Disparities in Incidence of COVID-19 Among Underrepresented Racial/Ethnic Groups in Counties Identified as Hotspots During June 5–18, 2020—22 States, February–June 2020. Morbidity and Mortality Weekly Report. 2020;69(33):1122.
24. Wortham JM. Characteristics of persons who died with COVID-19—United States, February 12–May 18, 2020. MMWR Morb Mortal Wkly Rep. 2020;69.
25. Fairlie RW, Couch K, Xu H. The Impacts of COVID-19 on Minority Unemployment: First Evidence from April 2020 CPS Microdata. National Bureau of Economic Research Working Paper Series. 2020;No. 27246.
26. Krieger N. ENOUGH: COVID-19, Structural Racism, Police Brutality, Plutocracy, Climate Change—and Time for Health Justice, Democratic Governance, and an Equitable, Sustainable Future. American Public Health Association; 2020.
27. Franco-Paredes C, Jankousky K, Schultz J, Bernfeld J, Cullen K, Quan NG, et al. COVID-19 in jails and prisons: A neglected infection in a marginalized population. PLOS Neglected Tropical Diseases. 2020;14(6):e0008409.
28. Saloner B, Parish K, Ward JA, DiLaura G, Dolovich S. COVID-19 Cases and Deaths in Federal and State Prisons. JAMA. 2020;324(6):602-3.
29. Chin T, Kahn R, Li R, Chen JT, Krieger N, Buckee CO, et al. US county-level characteristics to inform equitable COVID-19 response. MedRxiv. 2020.
30. Emeruwa UN, Ona S, Shaman JL, Turitz A, Wright JD, Gyamfi-Bannerman C, et al. Associations Between Built Environment, Neighborhood Socioeconomic Status, and SARS-CoV-2 Infection Among Pregnant Women in New York City. JAMA. 2020;324(4):390-2.
31. Syed ST, Gerber BS, Sharp LK. Traveling Towards Disease: Transportation Barriers to Health Care Access. Journal of Community Health. 2013;38(5):976-93.
32. VoPham T, Weaver MD, Adamkiewicz G, Hart JE. Social Distancing Associations with COVID-19 Infection and Mortality Are Modified by Crowding and Socioeconomic Status. Available at SSRN 3666297. 2020.
33. Karaye IM, Horney JA. The Impact of Social Vulnerability on COVID-19 in the U.S.: An Analysis of Spatially Varying Relationships. American Journal of Preventive Medicine. 2020;59(3):317-25.
34. Mikolai J, Keenan K, Kulu H. Intersecting household level health and socio-economic vulnerabilities and the COVID-19 crisis: An analysis from the UK. SSM - Population Health. 2020:100628.
35. Garg S. Hospitalization rates and characteristics of patients hospitalized with laboratory-confirmed coronavirus disease 2019—COVID-NET, 14 States, March 1–30, 2020. MMWR Morb Mortal Wkly Rep. 2020;69.

36. Williamson EJ, Walker AJ, Bhaskaran K, Bacon S, Bates C, Morton CE, et al. Factors associated with COVID-19-related death using OpenSAFELY. Nature. 2020;584(7821):430-6.
37. Zheng Z, Peng F, Xu B, Zhao J, Liu H, Peng J, et al. Risk factors of critical & mortal COVID-19 cases: A systematic literature review and meta-analysis. J Infect. 2020;81(2):e16-e25.
38. Zhou F, Yu T, Du R, Fan G, Liu Y, Liu Z, et al. Clinical course and risk factors for mortality of adult inpatients with COVID-19 in Wuhan, China: a retrospective cohort study. The Lancet. 2020;395(10229):1054-62.
39. Boyle CA, Fox MH, Havercamp SM, Zubler J. The public health response to the COVID-19 pandemic for people with disabilities. Disability and Health Journal. 2020;13(3):100943.
40. Turk MA, Landes SD, Formica MK, Goss KD. Intellectual and developmental disability and COVID-19 case-fatality trends: TriNetX analysis. Disability and Health Journal. 2020;13(3):100942.
41. Murray CJL, Lopez AD, Chin B, Feehan D, Hill KH. Estimation of potential global pandemic influenza mortality on the basis of vital registry data from the 1918–20 pandemic: a quantitative analysis. The Lancet. 2006;368(9554):2211-8.
42. Stevenson E, Barrios L, Cordell R, Delozier D, Gorman S, Koenig LJ, et al. Pandemic influenza planning: addressing the needs of children. Am J Public Health. 2009;99 Suppl 2(Suppl 2):S255-S60.
43. Kim L, Whitaker M, O'Halloran A, Kambhampati A, Chai SJ, Reingold A, et al. Hospitalization rates and characteristics of children aged< 18 years hospitalized with laboratory-confirmed COVID-19—COVID-NET, 14 states, March 1–July 25, 2020. Morbidity and Mortality Weekly Report. 2020;69(32):1081.
44. Fadini GP, Morieri ML, Longato E, Avogaro A. Prevalence and impact of diabetes among people infected with SARS-CoV-2. J Endocrinol Invest. 2020;43(6):867-9.
45. Pranata R, Lim MA, Huang I, Raharjo SB, Lukito AA. Hypertension is associated with increased mortality and severity of disease in COVID-19 pneumonia: A systematic review, meta-analysis and meta-regression. Journal of the renin-angiotensin-aldosterone system : JRAAS. 2020;21(2):1470320320926899.
46. Richardson S, Hirsch JS, Narasimhan M, Crawford JM, McGinn T, Davidson KW, et al. Presenting Characteristics, Comorbidities, and Outcomes Among 5700 Patients Hospitalized With COVID-19 in the New York City Area. JAMA. 2020;323(20):2052-9.
47. Yang J, Zheng Y, Gou X, Pu K, Chen Z, Guo Q, et al. Prevalence of comorbidities and its effects in patients infected with SARS-CoV-2: a systematic review and meta-analysis. International Journal of Infectious Diseases. 2020;94:91-5.
48. Curigliano G. Cancer Patients and Risk of Mortality for COVID-19. Cancer Cell. 2020;38(2):161-3.
49. Fung M, Babik JM. COVID-19 in Immunocompromised Hosts: What We Know So Far. Clinical Infectious Diseases. 2020.
50. Gao Y, Chen Y, Liu M, Shi S, Tian J. Impacts of immunosuppression and immunodeficiency on COVID-19: A systematic review and meta-analysis. J Infect. 2020;81(2):e93-e5.
51. Desmet K, Wacziarg R. Understanding Spatial Variation in COVID-19 across the United States. National Bureau of Economic Research; 2020. Report No.: 0898-2937.
52. Schuchat A. Public health response to the initiation and spread of pandemic COVID-19 in the United States, February 24–April 21, 2020. MMWR Morb Mortal Wkly Rep. 2020;69.

53. Zhang CH, Schwartz GG. Spatial Disparities in Coronavirus Incidence and Mortality in the United States: An Ecological Analysis as of May 2020. J Rural Health. 2020;36(3):433-45.
54. Cavallo JJ, Donoho DA, Forman HP. Hospital Capacity and Operations in the Coronavirus Disease 2019 (COVID-19) Pandemic—Planning for the Nth Patient. JAMA Health Forum. 2020;1(3):e200345-e.
55. Tsai TC, Jacobson BH, Jha AK. American hospital capacity and projected need for COVID-19 patient care. Health Affairs Blog. 2020.
56. Ku BS, Druss BG. Associations Between Primary Care Provider Shortage Areas and County-Level COVID-19 Infection and Mortality Rates in the USA. Journal of General Internal Medicine. 2020.
57. Epidemiology is a science of high importance. Nature Communications. 2018;9(1):1703.
58. Austin JM, Kachalia A. The State of Health Care Quality Measurement in the Era of COVID-19: The Importance of Doing Better. JAMA. 2020;324(4):333-4.
59. Liu Q, Luo D, Haase JE, Guo Q, Wang XQ, Liu S, et al. The experiences of health-care providers during the COVID-19 crisis in China: a qualitative study. The Lancet Global Health. 2020;8(6):e790-e8.
60. Center for Preparedness and Response, National Center for Environmental Health. Public Health Emergency Preparedness (PHEP) Cooperative Agreement: Centers for Disease Control and Prevention; [updated October 6, 2020. Available from: https://www.cdc.gov/cpr/readiness/phep.htm.
61. Times TNY. Nearly One-Third of U.S. Coronavirus Deaths Are Linked to Nursing Homes. New York Times. 2021 April 28, 2021.
62. USDA. The Meatpacking Industry in Rural America During the COVID-19 Pandemic 2020 [Available from: https://www.ers.usda.gov/covid-19/rural-america/meatpacking-industry/.
63. Development NCfO. O*NET OnLine 2020 [Available from: https://www.onetonline.org/.
64. Hamidi S, Sabouri S, Ewing R. Does density aggravate the COVID-19 pandemic? Early findings and lessons for planners. Journal of the American Planning Association. 2020;86(4):495-509.
65. Rufat S, Tate E, Emrich CT, Antolini F. How valid are social vulnerability models? Annals of the American Association of Geographers. 2019;109(4):1131-53.
66. Moore M, Gelfeld B, Okunogbe AT, Paul C. Identifying Future Disease Hot Spots: Infectious Disease Vulnerability Index: RAND Corporation; 2016.
67. Nguefack-Tsague G, Klasen S, Zucchini W. On Weighting the Components of the Human Development Index: A Statistical Justification. Journal of Human Development and Capabilities. 2011;12:183 - 202.
68. Wehrmeister F, Restrepo-Mendez M-C, França GV, Victora C, Barros A. Summary indices for monitoring universal coverage in maternal and child health care. Bulletin of the World Health Organization. 2016;94 12:903-12.
69. Burse N, Thompson E, Monger M. The Role of Public Health in COVID-19 Emergency Response Efforts From a Rural Health Perspective. Prev Chronic Dis. 2020;17.
70. Centers for Disease Control and Prevention. COVID-19 Secondary data and statistics 2020 [updated May 6. Available from: https://www.cdc.gov/library/researchguides/2019novelcoronavirus/datastatistics.html.
71. National Academies of Sciences E, Medicine. Framework for Equitable Allocation of COVID-19 Vaccine. Gayle H, Foege W, Brown L, Kahn B, editors. Washington, DC: The National Academies Press; 2020. 260 p.
72. Allen D, Bassuk A, Block S, Busenberg G, Charpignon M-L, Cohen J, et al. Pandemic Resilience: Getting it Done. COVID-19 Rapid Response Impact Initiative2020.

73. Siddarth D, Charpignon M-L, Foster D, Hoshino K, Kakade S, Langford JC, et al. Mitigate/Suppress/Maintain: Local Targets for Victory Over COVID. COVID-19 Rapid Response Impact Initiative2020.
74. World Food Programme. Nepal COVID-19 Economic Vulnerability Index. 2020 July 13.
75. Acharya R, Porwal A. A vulnerability index for the management of and response to the COVID-19 epidemic in India: an ecological study. The Lancet Global Health. 2020;8(9):e1142-e51.
76. The COVID mental health crisis in America's most vulnerable communities. Surgo Foundation and Mental Health America
2020 October 5, 2020.
